# Left ventricular trabecular layer operates at high ejection fraction: implications for pump function assessment in excessive trabeculation

**DOI:** 10.1101/2024.01.02.24300719

**Authors:** Ionela Simona Visoiu, Roxana Cristina Rimbas, Alina Ioana Nicula, Dragos Vinereanu, Bjarke Jensen

## Abstract

**Aims:** Numerous diagnostic criteria for excessive trabeculation (ET), or so-called “noncompaction”, score the extent of the trabecular layer, yet whether the trabeculations themselves have a negative or positive impact on pump function is largely unknown. This study aimed to measure the ejection fraction (EF) of the trabecular layer and its impact on pump function assessment.

**Methods and results:** We retrospectively analyzed cardiac magnetic resonance (CMR) findings in patients with ET of the left ventricle (LV). The LV was labelled into four regions: compact wall, central cavity (CC), trabeculations, and intertrabecular recesses (IR). For each label we calculated the systolic fractional volume change (SFVC) in short-axis images (n=15) and systolic fractional area change (SFAC) in 4-chamber images (n=30), by dividing end-systolic to end-diastolic values. We measured the EF of IR, CC, and total cavity (TC). Three methods to calculate EF of the TC were compared: trabeculations included (per guidelines), IR excluded (Jacquier criterion), trabeculations contoured and excluded (contour-EF).

The SFVC and SFAC of the compact wall were similar with SFVC and SFAC of trabeculations (both P>0.05). In contrast, the IR were more diminished in systole by comparison with the CC, having lower SFVC (39±17% vs. 56±16%, P<0.001) and SFAC (37±22% vs. 72±12%, P<0.001). EF of the IR was also greater than EF of the CC (61±17% vs. 44±16%, P<0.001). Excluding IR from the TC or including trabeculations underestimates the contour-EF (44±16% and 40±12%, respectively, vs. 51±16%; both P<0.001).

**Conclusions:** The trabecular layer has a high EF. Values of key prognostic indicators are better when this is accounted for.

**Graphical Abstract:** 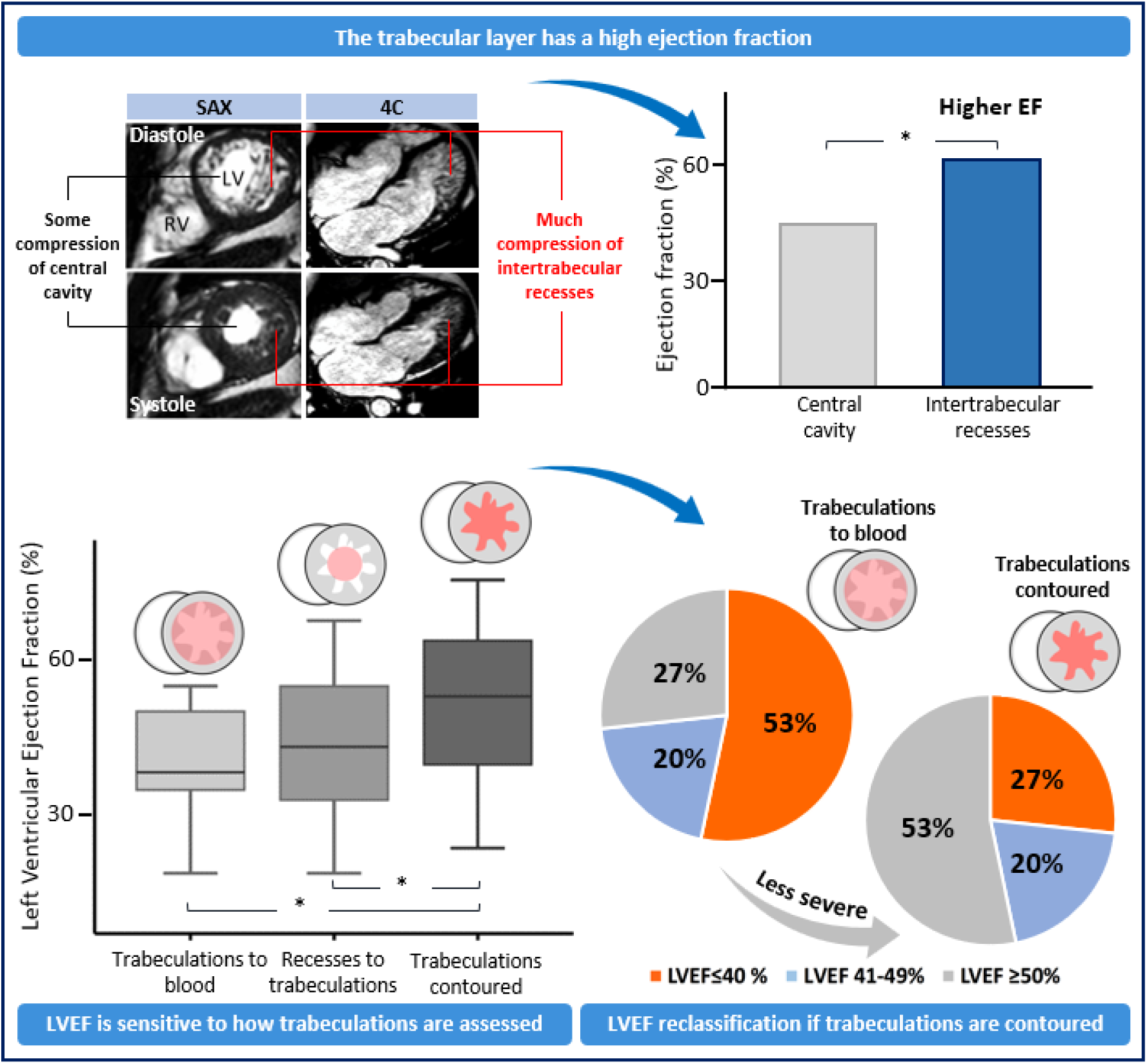

EF, ejection fraction; LVEF, left ventricular ejection fraction; SAX, short-axis; 4c, 4 chamber. *P<0001

## Introduction

Key prognostic indicators of heart function, such as left ventricle (LV) cardiac output, stroke volume (SV), ejection fraction (EF), and end diastolic volume (EDV) are currently measured by echocardiography or cardiac magnetic resonance imaging (CMR).^1^ Subtle variations in LV structure and function are now correlated to differences in quality of life and incidence of major adverse outcomes, as suggested by the big data analyses.^2–4^ One concern, however, is whether the functional readouts, which are often key prognostic indicators, are measured with the same accuracy across the various ventricular anatomies that the clinician encounters.

Imaging biases are consciously accepted in many instances even if the impact of them is only partly understood (*Figure 1*). The highly complex boundary between LV blood and trabeculations, for example, cannot be fully recognized with the spatial resolution of conventional clinical imaging.^5,6^ In echocardiography guidelines, papillary muscles and trabeculations are consciously neglected by adding them to the LV blood pool,^7^ and this is also the case in some CMR studies.^8^ While this is a pragmatic approach that strives to achieve highly reproducible assessments, it introduces a bias because the un-ejectable trabecular tissue is measured as blood (*Figure 1A*). A different bias is introduced when the intertrabecular recesses, which hold LV cavity blood, are added to the myocardial mass (*Figure 1B*).^9–12^ Attempts to avoid such biases have been made by contouring the trabeculations to the ventricular mass and the intertrabecular recesses to the LV blood pool (*Figure 1C*).^13–16^ These methods are in effect the application of a threshold value that is intermediate to the signal intensities of the compact wall and the blood of the central cavity. To what extent such thresholding is accurate is difficult to establish,^17^ because the thresholding is often performed on the images that are themselves considered the golden standard of cardiac imaging. A method of independent validation of the CMR-based contouring of the trabeculations is not commonly applied.

**Figure 1.**
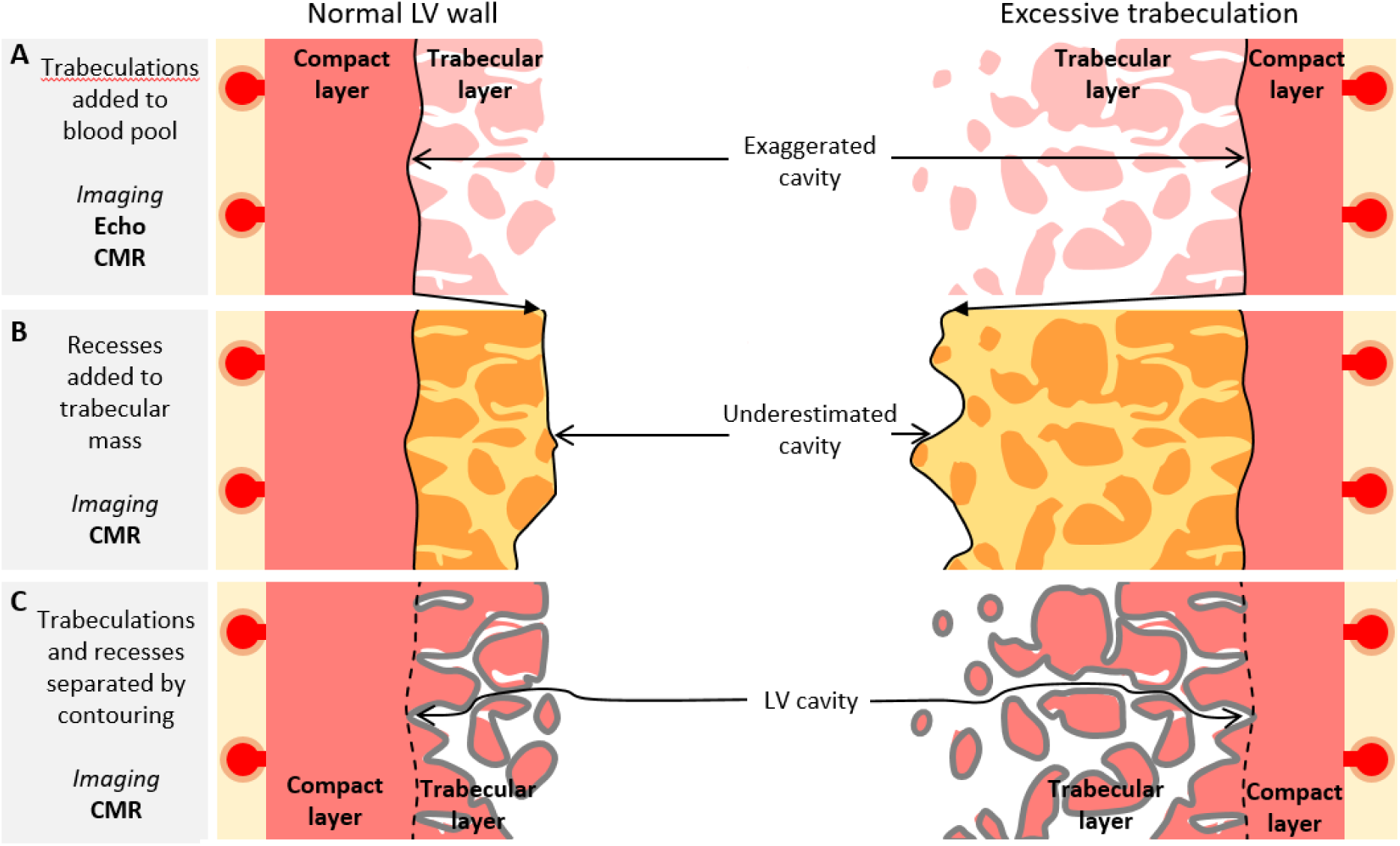
Schematic illustrations of biases in assessing LV volumes. **A**. Per guidelines, the trabeculations can be included in the cavity which then exaggerates the LV blood volume. **B**. Some quantifications of the trabecular mass (orange area) include the intertrabecular recesses, whereby the trabecular mass is exaggerated and LV blood volume is underestimated. **C**. Contouring (grey lines) separates blood and trabeculations, but its accuracy has no independent validation. Error in contouring, which is normal, is illustrated by imperfect lining of the trabecular contours.

The methodological biases in measuring key prognostic indicators can be expected to be greater with the degree of LV trabeculation, being exaggerated in individuals with excessive trabeculation (ET), also known as noncompaction, or hypertrabeculation, or persistent sinusoids (*Figure 1*).^18^ We hypothesized that the trabecular layer operates at higher EF than the central cavity. This study focuses on individuals with ET since their trabecular layer is larger than normal and the EF will be correspondingly easier to measure.

## Methods

### Study design and population

We conducted a retrospective study on patients evaluated by CMR and diagnosed with ET in one center (University and Emergency Hospital, Bucharest, Romania). The inclusion criteria were: positive Jacquier criterion^11^ (the trabecular myocardium comprises more than 20% of the LV mass), presence of sinus rhythm, and age over 18 years. We excluded patients with chronic coronary syndromes, significant valvular heart diseases, pericardial diseases, or poor quality of CMR images. 15 patients were included in the study (*Table 1*). The study was approved by the Research Ethics Committee of the University and Emergency Hospital (Bucharest, Romania) (approval number: 74155), and complied with the Declaration of Helsinki. All participants provided informed written consent.

**Table 1.** Characteristics of the study population.

|  | ET patients (n=15) |
| --- | --- |
| Age (years) | 48.3 ± 12.5 |
| Male (%) | 53.3 |
| BMI (kg/m<sup>2</sup>) | 29.5 ± 5.9 |
| Systolic blood pressure (mmHg) | 135.9 ± 25.5 |
| Diastolic blood pressure (mmHg) | 81.7 ± 14.4 |
| Trabeculations % of total LV myocardium | 36.3 ± 8.3 |
| Trabeculations volume (ml) | 37.0 (33.5-70.5) |
| Trabecular layer volume (ml) | 130.0 ± 48.9 |
| Trabeculations % of the trabecular layer | 37.3 ± 7.4 |

Continuous variables are presented as mean ± SD or median (IQR); BMI, body mass index; LV, left ventricle.

### Cardiac magnetic resonance

CMR was performed using a 1.5-T MR scanner (Magnetom Sempra, Siemens Healthcare GmnH, Erlanger, Germany). Standard short- and long-axis cine images were acquired using steady-state free precession sequences. Frames of end-diastole (ED) and end-systole (ES) were exported to and analyzed in the 3D software Amira (version 3D 2021.2, FEI SAS, Thermo Fisher Scientific), where all structures were labelled in the Segmentation Editor module. The volumes of labelled structures were retrieved using the Materials Statistics module. To differentiate myocardium from lumen, a mask/signal threshold was applied (per heart) to the ED 4-chamber frame (long-axis) or a mid-ventricular frame (short-axis stack) such that the papillary muscles were myocardial and the trabecular layer became approximately equal parts trabeculations and intertrabecular recesses. Then the threshold was locked and applied to all other ED and ES frames of the images series of that heart. Thus, the LV was labelled into four regions in both ED and ES: compact wall, central cavity, trabeculations, and intertrabecular recesses.

Volumes were measured on the basis of the short-axis stacks, while areas on the basis of 4-chamber images. For each label we calculated the systolic fractional volume change (SFVC) and systolic fractional area change (SFAC), by dividing ES to ED values. The volumes of trabeculations and compact wall were calculated as the average of the volume measured in ED and ES. From the labels of central cavity and intertrabecular recesses we calculated EDV, end-systolic volume (ESV), SV, and EF. This was done in the three different ways illustrated in *Figure 1*. In the first way, according to the current guidelines,^7^ the total LV cavity also comprised trabecular myocardium besides the central cavity and the intertrabecular recesses (*Figure 1A*). In the second way, according to Jacquier et al.,^11^ the total LV cavity excluded the intertrabecular recesses, which were considered trabecular myocardium (*Figure 1B*). In the third way, or ‘contour’, which is the reference in this study, the intertrabecular recesses were considered part of the total LV cavity (*Figure 1C*).

Using syngo.MR Cardiology VB20A post-processing software (syngo.via, Siemens Healthcare GmnH, Erlanger, Germany) we measured mitral annular plane systolic excursion (MAPSE) and LV length. MAPSE was calculated as the difference between ED to ES of the wall length, measured as a straight line from epicardial apex to the mitral annulus. We measured MAPSE for all six walls from 2-chamber, 3-chamber, and 4-chamber cine images. We calculated the global MAPSE as the average of all six measurements. LV length was defined as the distance from the epicardial apex and the midpoint of the line connecting the origins of the mitral leaflets. LV length was measured in ED and ES in all three standard apical views. The global LV length was calculated as the average of the three measurements. Based on LV length we calculated the global longitudinal shortening (GL-shortening), according to the bellow formula, as previously reported:^19^ GL-shortening = 100 × (LV_length ED−LV_length ES) ∕ LV length ED.

### Analysis of published images

To test the robustness of our findings, we included 4-chamber views publicly available cases^20–29^ of excessive or abnormal trabeculation from https://radiopaedia.org and from publications.^18,30–33^ The inclusion criterion was available images of both ED and ES.

### Theoretical model

We used simple theoretical calculations to evaluate the effect of trabeculations on the ventricular performance, using the baseline values listed in *Table 2*. A normal LV was compared to one with ET, and in both cases the total LV cavity and trabeculations amounted to 150 ml. With the total volume clamped at 150 ml, we then calculated what happened to blood volumes if one parameter such as the EF of the intertrabecular recesses varied from 10 to 90%.

**Table 2.**
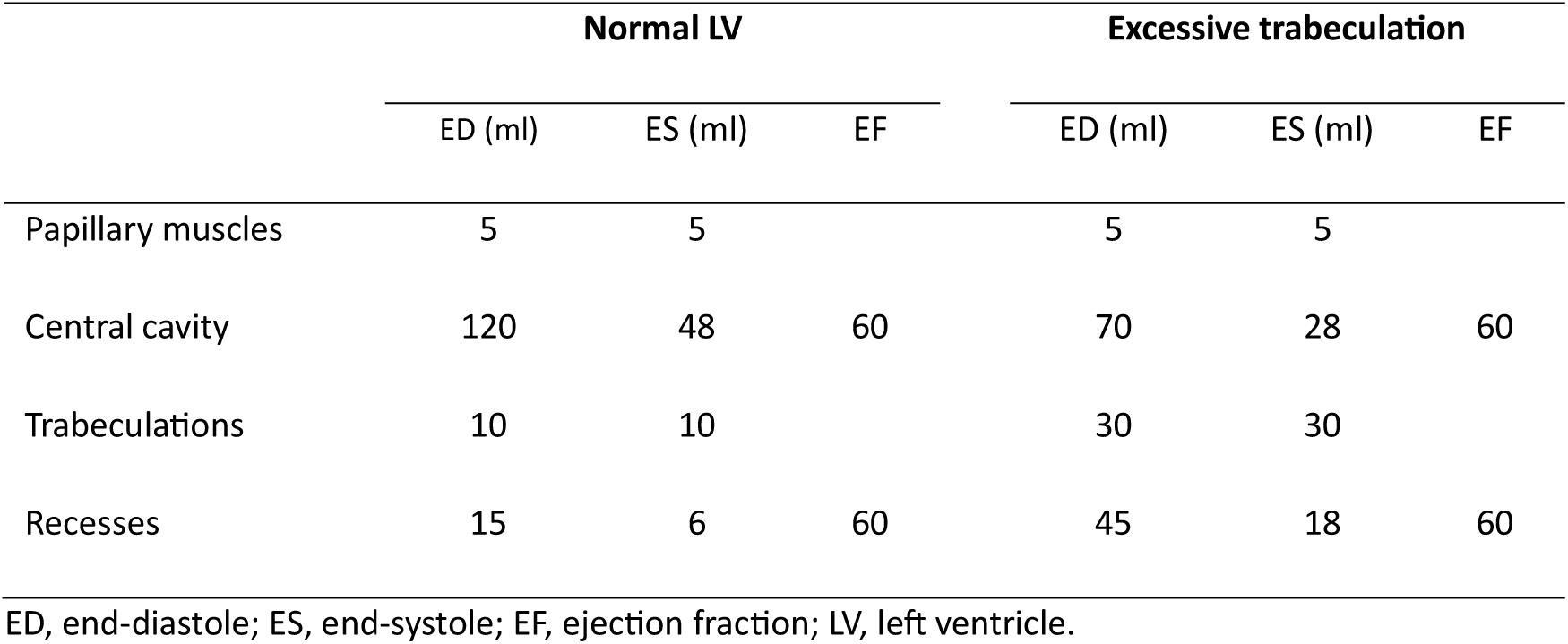
Baseline values for theoretical calculations.

### Statistical analysis

Statistical analysis was performed using the SPSS version 21.0 (IBM Corp., Armonk, NY, USA). Categorical variables were expressed as percentages. Continuous variables were assessed for normality using the Shapiro–Wilk and Kolmogorov-Smirnov tests. Normally distributed continuous variables were reported as mean ± standard deviation (SD), and were compared for statistical significance using one-sample and paired-samples t-tests. Non-normally distributed continuous variables were reported as median and interquartile range (IQR). Correlations between normally distributed continuous variables were performed using Pearson’s correlation coefficient. A p-value < 0.05 was considered statistically significant.

## Results

### Assessment of the trabecular layer based on short-axis views

First, we describe how the volume of the four left ventricular labels change from diastole to systole to document if the readouts of the labelling correspond to expected outcomes (*Figure 2A-B*). For each label, we measured the SFVC. Total left ventricular volume (tissue & blood) should be smaller in systole, and this was the case (*Figure 2C*). The volume of myocardium should not be different between diastole and systole, and this was the case for the compact layer, whereas the volume of trabeculations was slightly and significantly greater in systole (*Figure 2C*). The blood volume should be lower in systole and both the volume of the central cavity and the intertrabecular recesses were substantially lower in systole (*Figure 2C*). Then, we found that the volume of intertrabecular recesses was significantly more reduced by comparison with the central cavity (*Figure 2C*), having lower SFVC values (39±17 vs. 56±16%, P<0.001). When the same volume changes were calculated as EF (EDV-ESV/EDV), the EF of intertrabecular recesses was significantly greater than that of the central cavity (61±17 vs. 44±16%, P<0.001). It required very substantial mislabeling of the central cavity (deflation) and the recesses (inflation) to make the EF of intertrabecular recesses as low as the EF of the central cavity (Supplementary data online *Figure S1*). To further validate the labelling, we compared contour-EF of the total ventricle to GL-Shortening and MAPSE, the measurements of which can be made without consideration to the trabecular layer. The contour-EF was positively correlated to both GL-Shortening (R^2^=0.341, P=0.021) and MAPSE (R^2^=0.268, P=0.045) (Supplementary data online *Figure S2*).

**Figure 2.**
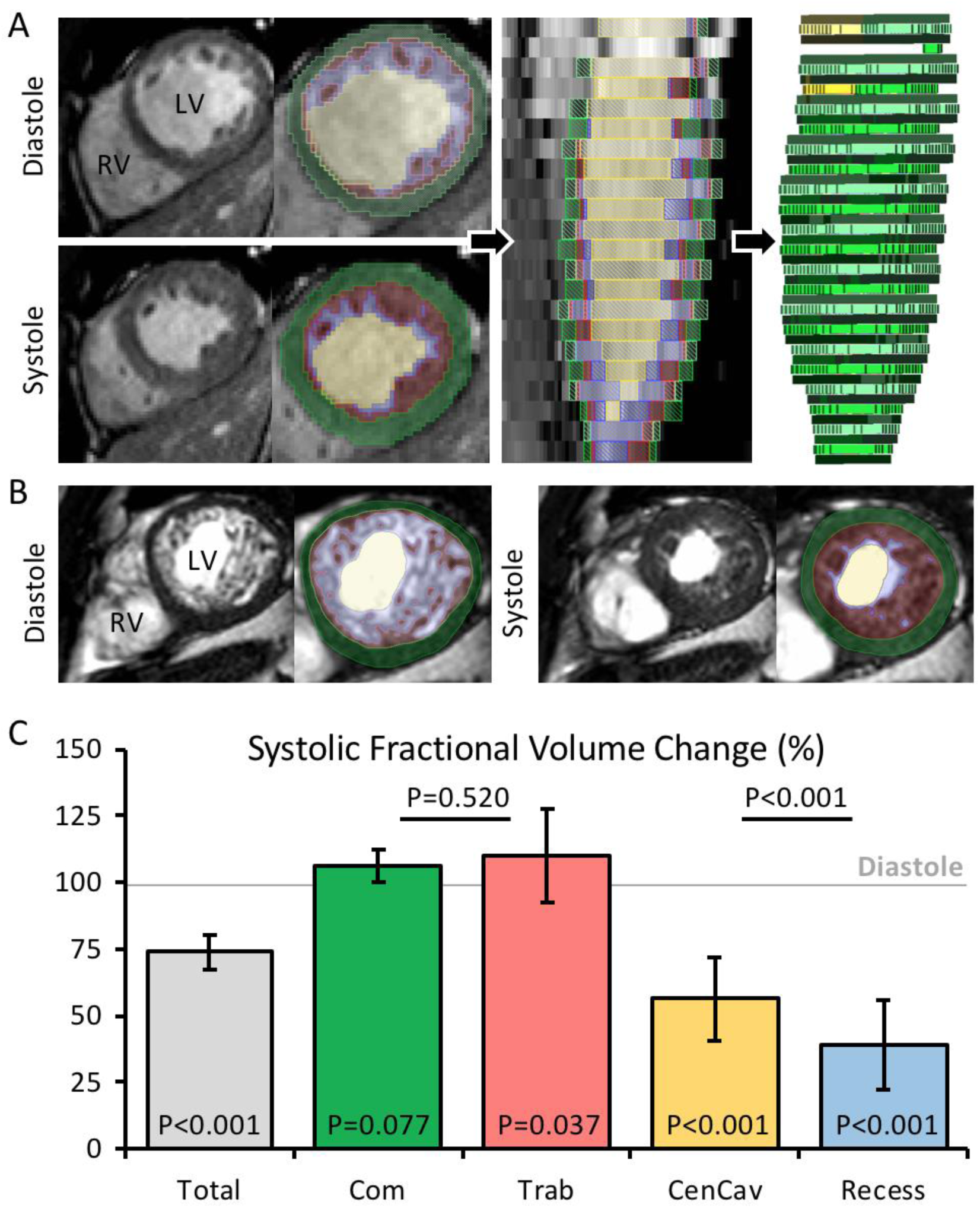
Volume changes of the four major components of the total left ventricle. **A**. Example of threshold-based labelling of a short-axis slice. **B**. Example of threshold-based labelling of a short-axis slice from the individual with the greatest proportion of LV trabecular myocardium. **C**. Volume change between diastole and systole for the sum of all labels (Total) and each of the four labels (Com, compact layer; Trab, trabeculations; CenCav, central cavity; Recess, intertrabecular recesses). The P-value within each column, is the one-sample t-test for difference from 1 (diastole). The P-values between columns are paired-sample t-tests for the same mean (N=15).

### Assessment of the trabecular layer based on 4-chamber views

Since LV morphology and function are often visualized in 4-chamber view, we made the same analysis for the areas of the four labels in single slice mid 4-chamber view (*Figure 3A*). First, we correlated the already measured SFVC of the central cavity and the recesses from the short-axis stacks, to the corresponding SFAC from 4-chamber views; area changes were positively correlated to volume changes for both labels (*Figure 3B*). Given this correlation, we expanded our dataset from 15 to 30 cases using publicly available cases. In systole, the total left ventricular area (tissue & blood) is likely to be smaller, and this was the case (*Figure 3C*). The area of myocardium is expected to be greater in systole, because the total myocardial volume will be distributed on fewer frames, and both the compact and trabecular layer labels had a greater area in systole (*Figure 3C*). The cavity areas should be lower in systole, if there is intermediate to good contraction, and this was the case, both for the central cavity and the intertrabecular recesses (*Figure 3C*). In correspondence to what was found with the volume measurements, the area reduction was significantly greater for intertrabecular recesses than for the central cavity (*Figure 3C*), having lower SFAC values (37±22% vs. 72±12%, P<0.001, paired t-test).

**Figure 3.**
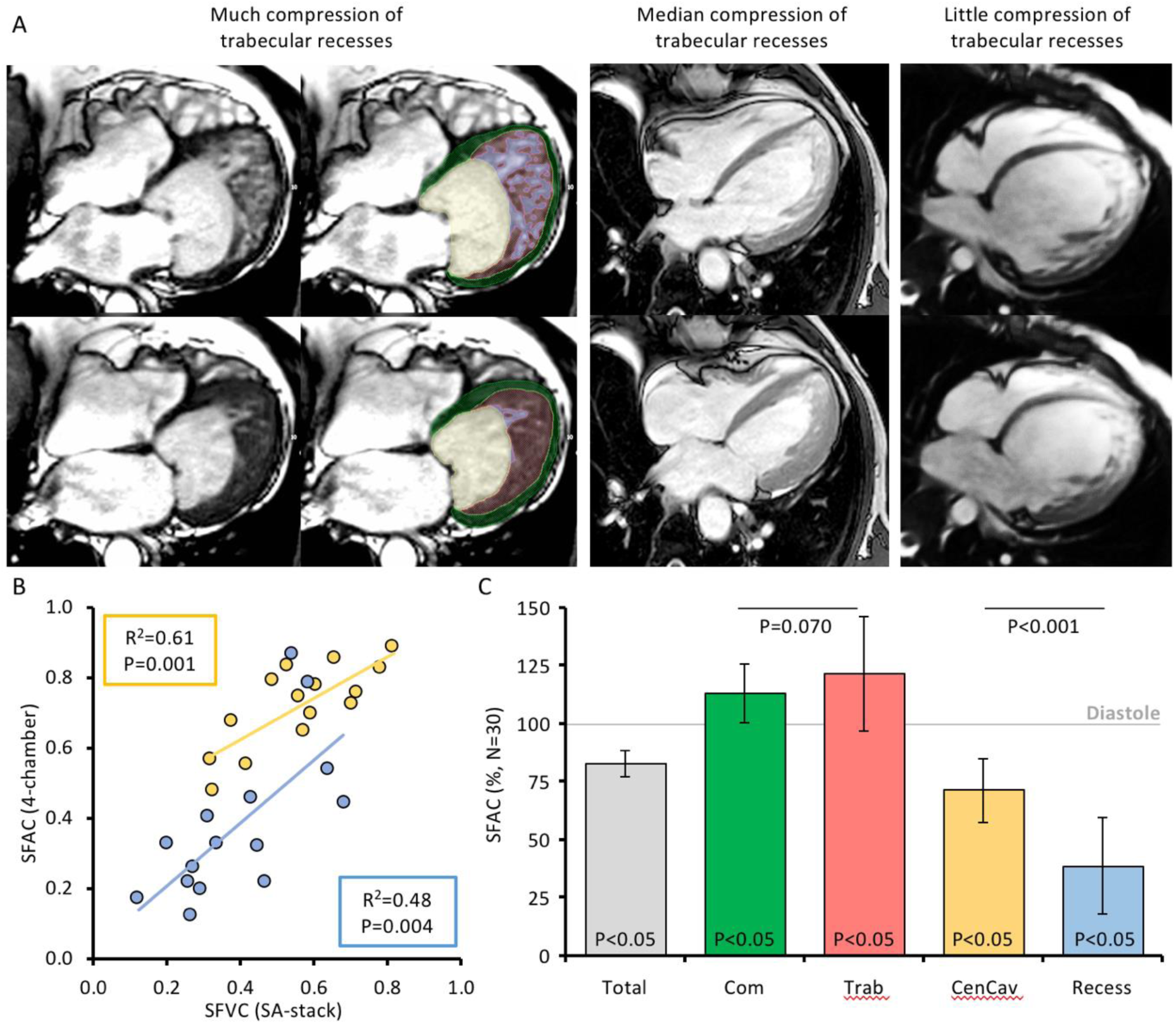
Area changes of the four major components of the left ventricle. **A**. Examples of left ventricles in diastole (top row) and systole (bottom row), in which the systolic intertrabecular recesses are much compressed (left-hand image - case from Petersen et al.^18^), intermediately compressed (middle image - case from Radiopaedia, Luikjx^24^), or show little compression (right-hand image - case from Radiopaedia, Yarmola^29^). **B**. There were significant linear correlations between area (SFAC) and volume (SFVC) changes, both for the central cavity (yellow) and the intertrabecular recesses (blue). **C**. Area change between diastole and systole for the sum of all labels (Total) and each of the four labels (Com, compact layer; Trab, trabeculations; CenCav, central cavity; Recess, intertrabecular recesses). The P-value within each column, is the one-sample t-test for difference from 1 (diastole). Both the compact and trabecular layer areas are significantly greater in systole than in diastole, but not different from each other. In contrast, the intertrabecular recess areas are more diminished in systole than is the central cavity.

### Impact of the trabecular layer on LV cavity volume assessments

Given the higher EF of the trabecular layer than the central cavity, we next expanded the analysis of our volumetric data to assess the impact of adding the volume of trabeculations to the blood, that is the combined volume of the central cavity and the recesses (per guidelines). Compared to the volumes derived from contoured trabeculations, EDV and ESV were increased (*Figure 4*). The SV was not different between the two methods, but given the increased EDV, EF was diminished (*Figure 4*). We then assessed the impact of excluding the intertrabecular recesses from the blood pool (per Jacquier criterion^11^). This was done by subtracting the intertrabecular recesses volume from LV cavity volume, and then we compare these volumes to the volumes derived from contoured trabeculations. All volumes and the EF were diminished (*Figure 4*).

**Figure 4.**
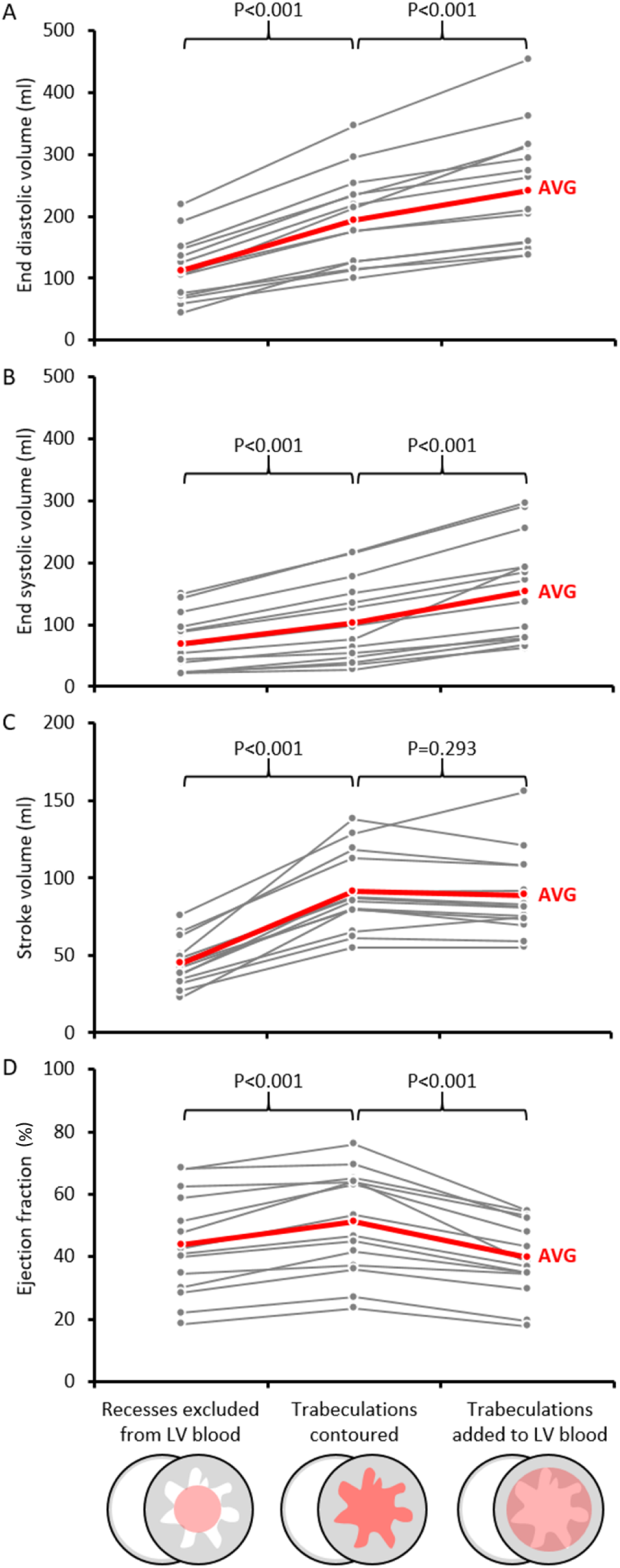
How differences in analyses of the trabecular layer impact on LV cavity volume assessments. **A**-**B**. Compared to trabeculations contouring, the end diastolic (A) and systolic (B) volumes are diminished if recesses are excluded from the blood pool (Jacquier criterion) and are exaggerated if trabeculations are added to the blood pool (per guidelines). **C**. The exclusion of the recesses has a negative impact on the stroke volume. **D**. The ejection fraction has the highest value if trabeculations are contoured.

### Calculations to show how trabecular layer characteristics can affect assessment of volumes

The above analyses suggest the specifics of trabecular layer characteristics, such as EF, can affect volume assessments. Next, we made theoretical permutations to key characteristics, the baseline values of which are listed in *Table 2*, to assess the scope of these effects. If trabeculations are included in the LV blood pool and the intertrabecular recesses and the central cavity both operate at an EF of 60%, the greater the trabecular tissue volume, the lower the measured EF will be (*Figure 5A,* Supplementary data online *Figure S3*). In failing hearts, the ventricular wall can have much subendocardial fibroelastosis and this can reduce the compliance of the trabecular layer.^26,34^ Varying the EF of an excessive trabecular layer from 10% (extremely reduced compliance) to 90%, while keeping the central cavity EF at 60%, shows the total EF to increase from below 40% to above 70% (*Figure 5B*, Supplementary data online *Figure S4*). Myocardial volume is considered easier to measure in systole^13^ and our own data show the volume measurements of the trabeculations can vary between diastole and systole. If the trabecular tissue volume is underestimated in diastole, it will lead to an overestimation of the EDV, SV, and EF (*Figure 5C*, Supplementary data online *Figure S5*).

**Figure 5.**
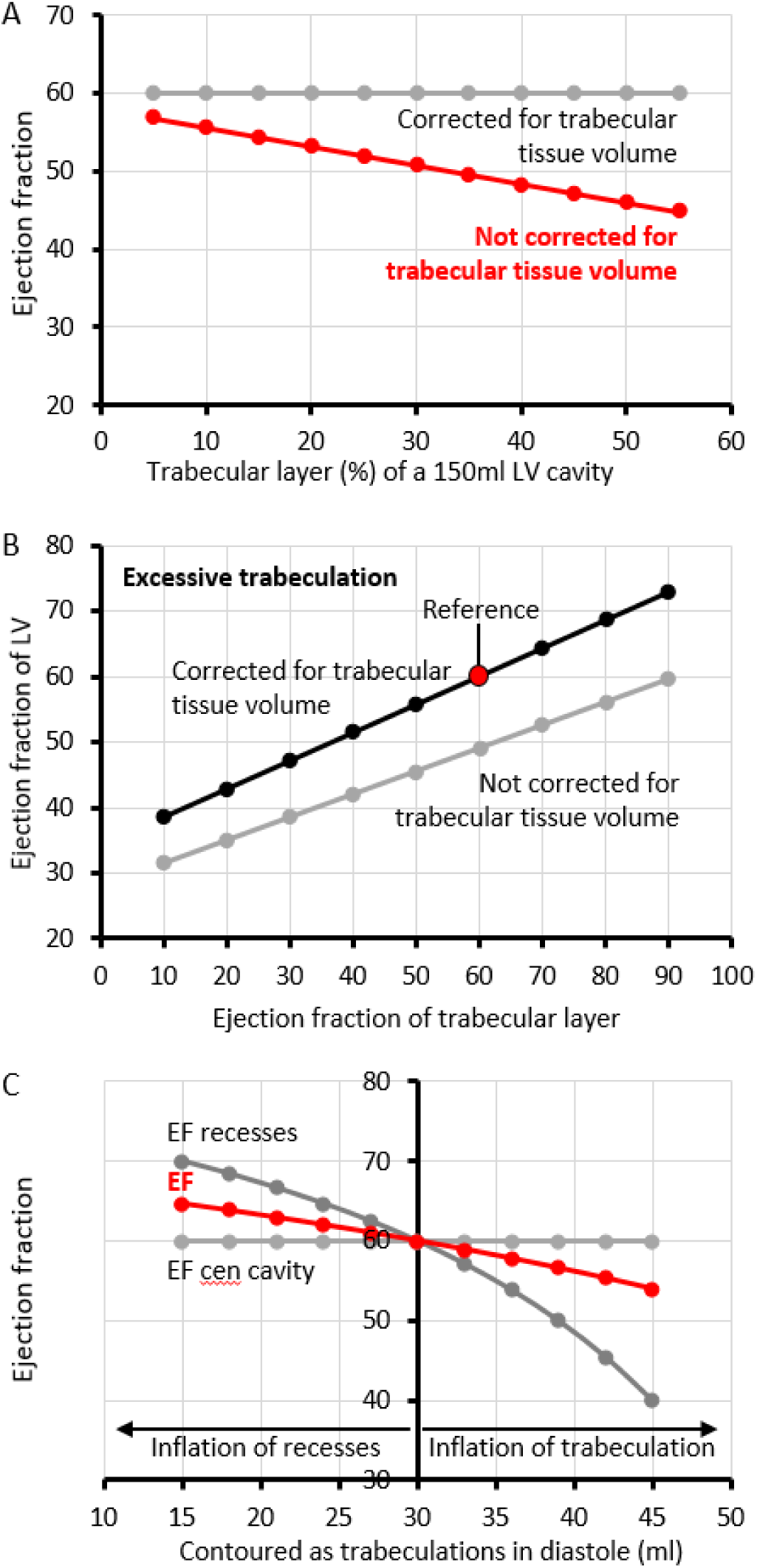
Theoretical permutations to key characteristics of the trabecular layer and its impact on ejection fraction. **A**. The greater the proportion of trabecular muscle the lower the ejection fraction, unless the trabecular myocardial volume is corrected for. **B**. The greater the ejection fraction of the trabecular layer, the greater the total ventricular ejection fraction. The ejection fraction of the central cavity was kept at 60%. **C**. The greater the diastolic measurement of trabecular myocardium deviates from the systolic measurement (30ml), the greater the perturbation to the ejection fraction of the intertrabecular recesses (EF) and the total LV ejection fraction (EF).

### LVEF category reclassification after contouring of trabeculations

So far, in the analyses performed, there was nothing to indicate that the trabecular proportion of total LV myocardium was predictive of the trabecular layer function. Next, we divided the patient population on the contour-EF of the total cavity being above or below 50%, and in both of the resultant groups, the EF of the recesses remained significantly greater than that of the central cavity (*Table 3*). Also, there was no significant linear correlation between total EF and the extent of trabeculation (P=0.471, *Figure 6A*). Finally, we classified patients based on contoured trabeculations and per guidelines, and the number of patients with severely reduced EF was halved if the EF was based on contouring of the trabeculations (*Figure 6B*).

**Figure 6.**
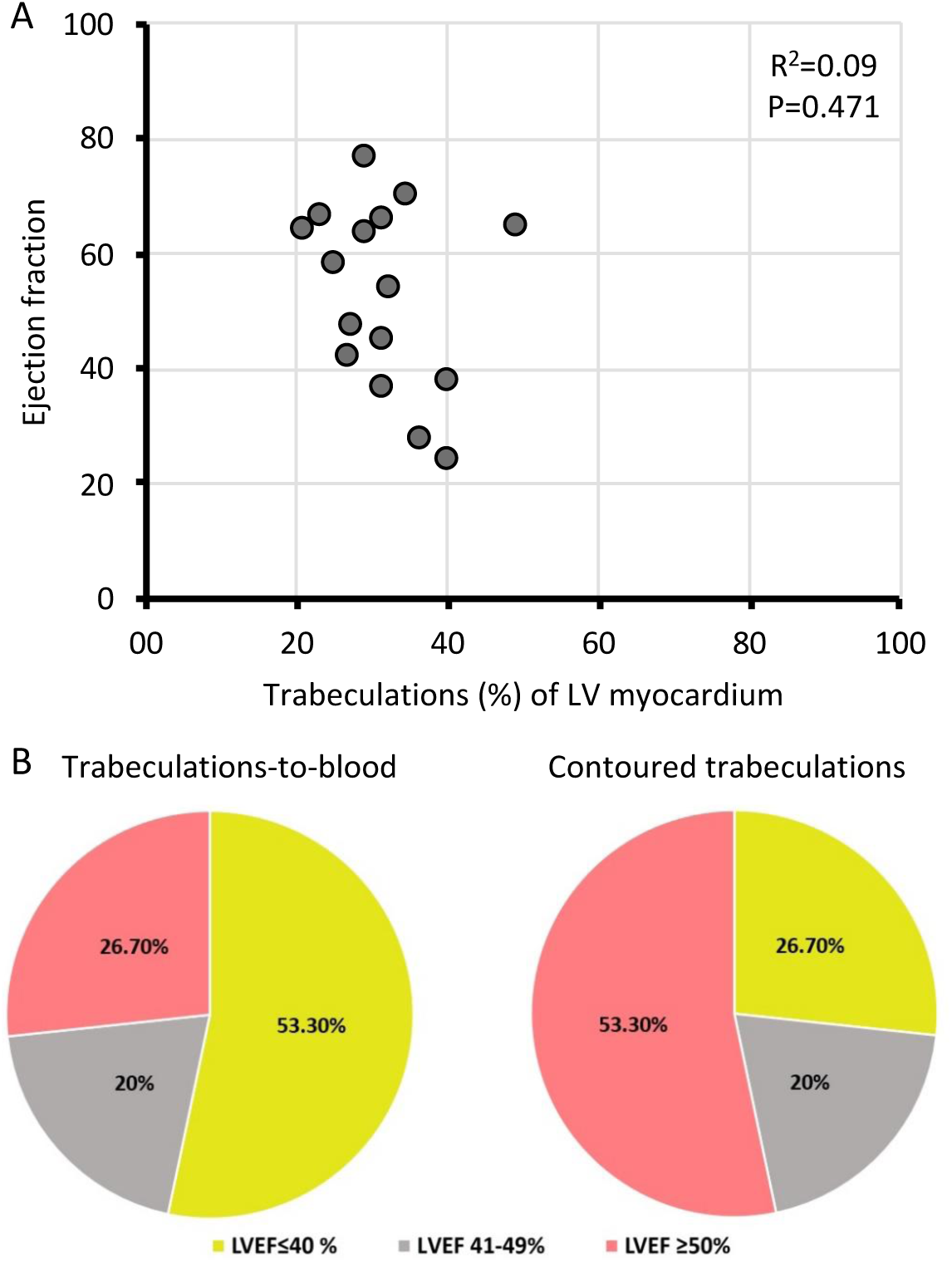
Impact of trabeculations on ejection fractions. **A**. When trabeculations were contoured, there was no significant correlation between total LV ejection fraction (LVEF) and the proportion of trabeculation. **B**. LVEF category reclassification after contouring of trabeculations.

**Table 3.** Comparison between the central cavity EF and recesses EF.

| Variable | All patients<br>(n=15) | LVEF<50%<br>(n=7) | LVEF>50%<br>(n=8) |
| --- | --- | --- | --- |
| Central cavity EF (%) | 43.8 ± 15.8 | 30.6 ± 8.7 | 55.4 ± 10.2 |
| Recesses EF (%) | 61.0 ± 16.7 | 45.9 ± 9.6 | 74.3 ± 6.9 |
| P value | <0.001 | <0.001 | <0.002 |

EF, ejection fraction; LVEF, left ventricular ejection fraction.

## Discussion

To the best of our knowledge this is the first study to analyze separately the intertrabecular recesses and the central cavity for the volume changes that occur between diastole and systole. The main finding is that the intertrabecular recesses operate at a high EF. This finding is consistent with the reproducible correlation between high levels of physical activity and a greater extent of the trabecular layer.^2,4^ Conversely, it undermines the notion that the trabecular layer has a direct negative impact on pump function. In the context of so-called noncompaction cardiomyopathy this notion is widespread, either explicitly or implicitly, but it is a notion that is becoming increasingly untenable.^18^ Part of the evidence against the notion is that the human ventricular trabecular and compact myocardium is not different in the density of sarcomeres, mitochondria, and vasculature,^35^ and single cell sequencing does not reveal overt differences between trabecular and compact myocardium.^36^ In our interpretation of the current literature, the trabecular and compact layers are made up of similar myocardium. One difference may be that the trabecular layer more readily can collapse on itself than the compact wall, i.e. it can achieve a very high EF.

We show that when ventricular volumes and EF are measured, there is a substantial impact of adding the trabeculations to the LV blood pool, as per guidelines^7^ or removing the intertrabecular recesses from the LV cavity, as per a criterion for noncompaction.^11^ These practices likely reflect pragmatic choices to create easily standardized methodologies, even if these practices introduce biases. The challenge is that it is difficult for the clinician to compute on-the-spot what these biases are, and derive how they affect key parameters. And these biases are exaggerated if the trabecular layer is greater than normal. We show, in accordance with previous studies on normal hearts,^14^ that contouring of the trabeculations impacts on volume measurements. Because we have based our investigations on individuals with ET, the effects of contouring are pronounced. In our population, our best contouring led to a halving of the population with an EF less than 40%, and it doubled the number of individuals with an EF greater than 50%. In effect, many of our current manners of assessing LV function bias towards reduced function when there is a setting of ET. These more accurate volume measurements may have also important clinical implications in patients with ET and heart failure (HF). Thus, half of the patients with an EF of less than 40% would no longer have an indication to initiate some of the four treatment pillars in HF, according to the current guidelines.^37^ It will also lead to a significant reduction in device therapies indications in patients with ET. Moreover, a higher EF than previously reported would explain the overall good prognosis in individuals with ET over 9.5 years follow-up.^38^

The trabecular layer, however, cannot be analyzed in isolation. Like the ventricular cavity, it is ultimately enclosed by the compact wall. We presume that the high EF of the intertrabecular recesses reflects the work done both by the trabecular layer and the compact wall. In addition, the high EF likely reflects the intertrabecular recesses eject into the central cavity, whereas the central cavity is a conduit. In systole, it will empty into the aorta while concurrently it will be filled with blood from the intertrabecular recesses. Therefore, the greater the trabecular layer, and the greater the systolic compression of the intertrabecular recesses, the greater the systolic filling will be of the central cavity. In this way, a greater trabecular layer biases towards a lower measured EF of the central cavity, even if the SV is in the normal range relative to the LV mass and volume. Interestingly, the trabecular layer may intrinsically allow for a greater EF than the compact wall, given that in the individuals with a total EF above 50%, their intertrabecular recesses operated with an average EF of 74%. Indeed, animals with highly trabeculated ventricles can achieve EF of approximately 90%.^39^

### Study limitations

The fractional area change and volume readouts reported here derive from manual labelling of the structures. Some challenges of this approach^15^ are overcome by the use of a threshold value to separate cavity from myocardium. Such threshold can be determined on a case-by-case basis since there are slight variations between individual recording sessions. We applied the same threshold in diastole and systole since it seemed the simplest approach to us, but this approach may not be the best. The use of thresholds is inadequate to separate compact from trabecular myocardium, and to separate the central cavity from intertrabecular recesses. For these distinctions, one relies on identification of landmarks, such as the depth of intertrabecular recesses, and ‘common sense’ assumptions, such as that the compact wall will take the shape of a ring of more or less even thickness. This manner of analysis, then, necessarily comes with some technical variation. The difference in EF of the intertrabecular recesses versus the central cavity, however, is so great that this finding is very resilient to variation labeling of structures, as we show in Supplementary data online *Figure S1*. Being a CMR imaging-focused study we did not include other clinical data. Because we selected patients with a positive Jacquier criterion, in order to analyse a well-defined trabecular layer, we identified a low number of patients with ET, from a single center, and this study could be considered hypothesis-generating research that needs validation in a large-scale study.

## Conclusions

The trabecular layer is associated with a high EF. By contouring the trabeculations, LV volumes are more accurately assessed, and implicitly the EF. The contour-EF, which actually is much higher, could better guide the clinical management in daily practice, avoiding inappropriate diagnosis and treatment. It may also better predict prognosis in patients with excessive trabeculation in future prospective studies.

## Data Availability

All data produced in the present study are available upon reasonable request to the corresponding author.

## SUPPLEMENTARY MATERIAL

**Supplementary Figure S1.**
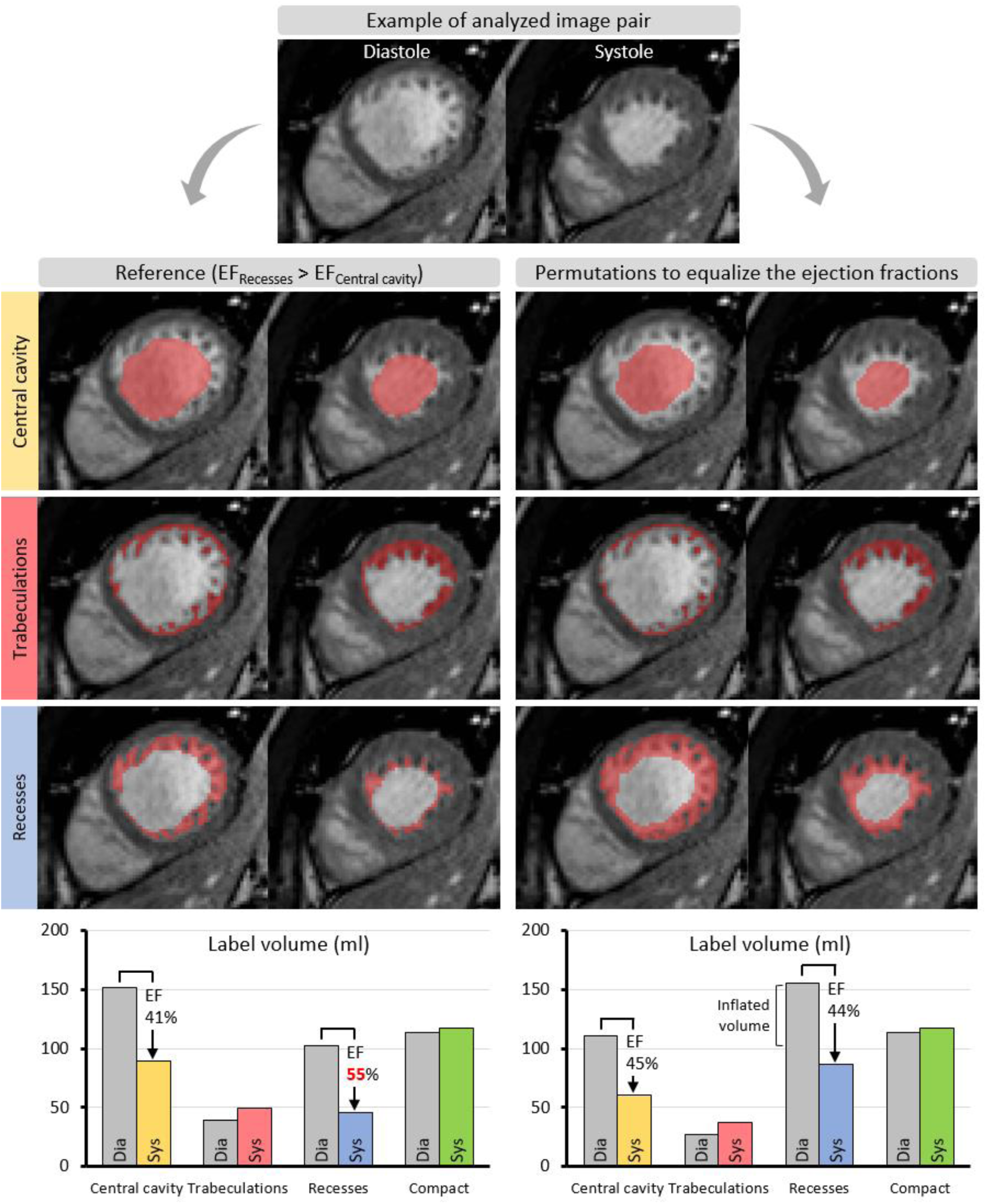
Very substantial mislabeling of the intertrabecular recesses (inflation) is required, at the cost of the central cavity and trabeculations labels, to lower the EF of the intertrabecular recesses to the EF of the central cavity (in this case from 55 to 44%).

**Supplementary Figure S2.**
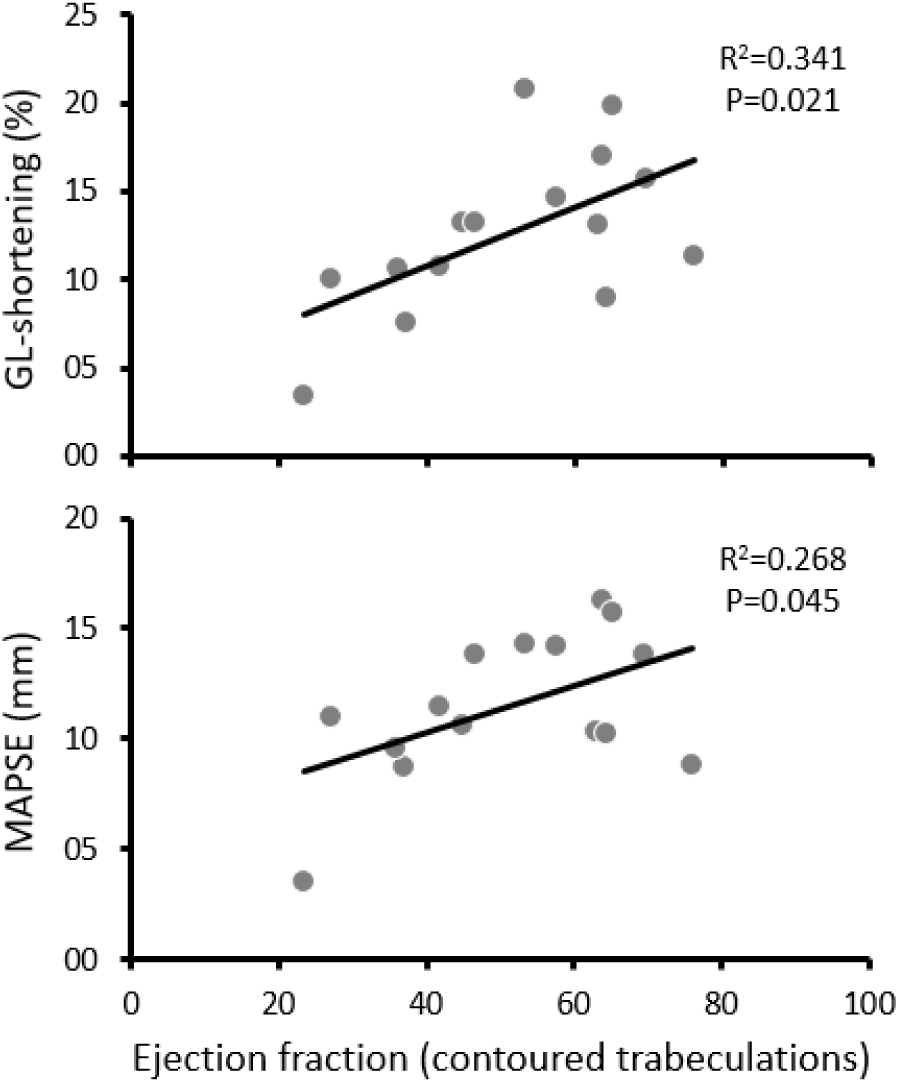
The ejection fraction derived from frames with contoured trabeculations was significantly correlated to both global longitudinal shortening (GL-Shortening) and mitral annular plane systolic excursion (MAPSE). P-values relate to linear regressions.

**Supplementary Figure S3.**
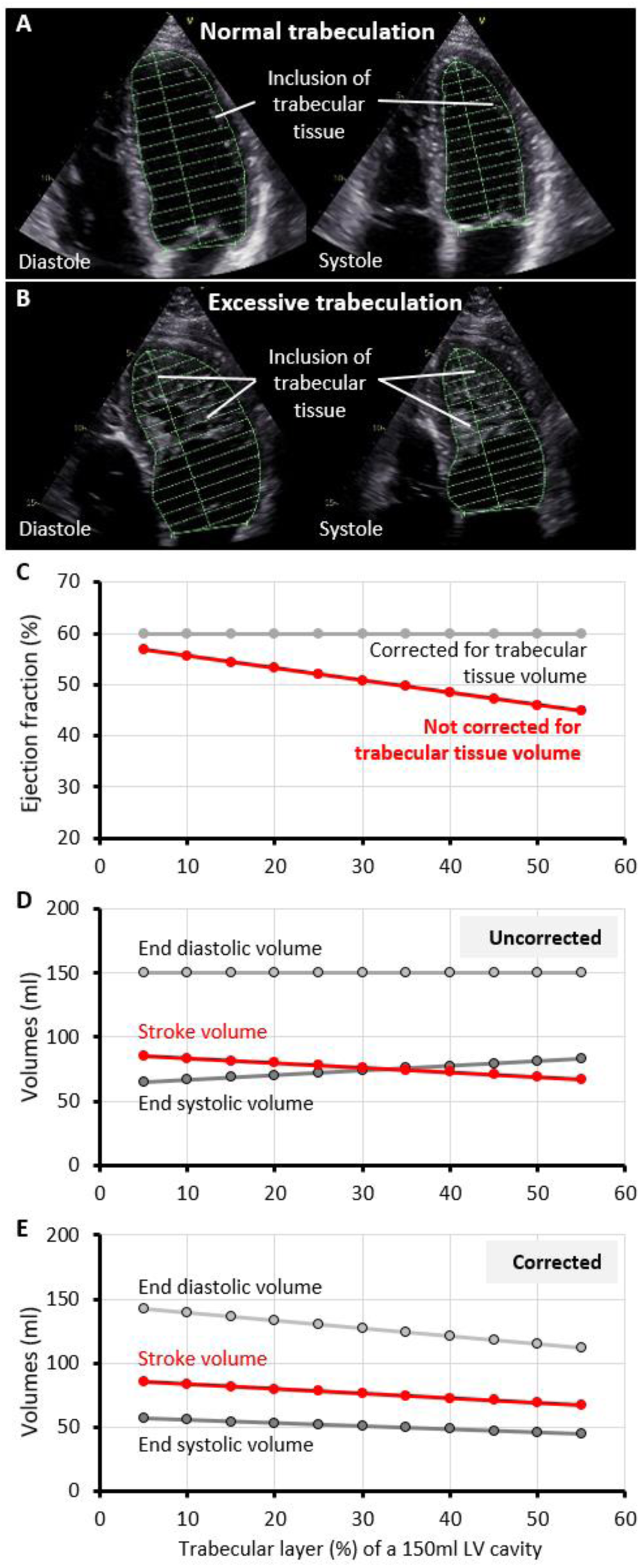
Impact of including trabeculations in the LV blood pool. **A**-**B**. Echocardiography of a left ventricle with a normal extent of trabeculations (**A**) and with excessive trabeculations (**B**). **C**. The greater the proportion of the trabecular layer, the greater the reduction in ejection fraction. **D**. More trabeculations equates more un-ejectable tissue in the cavity as define per guidelines (Lang et al), which increases the end systolic volume and thus diminishes the stroke volume. **E**. If the un-ejectable trabeculations are corrected for by accurate contouring, all volumes diminish isometrically.

**Supplementary Figure S4.**
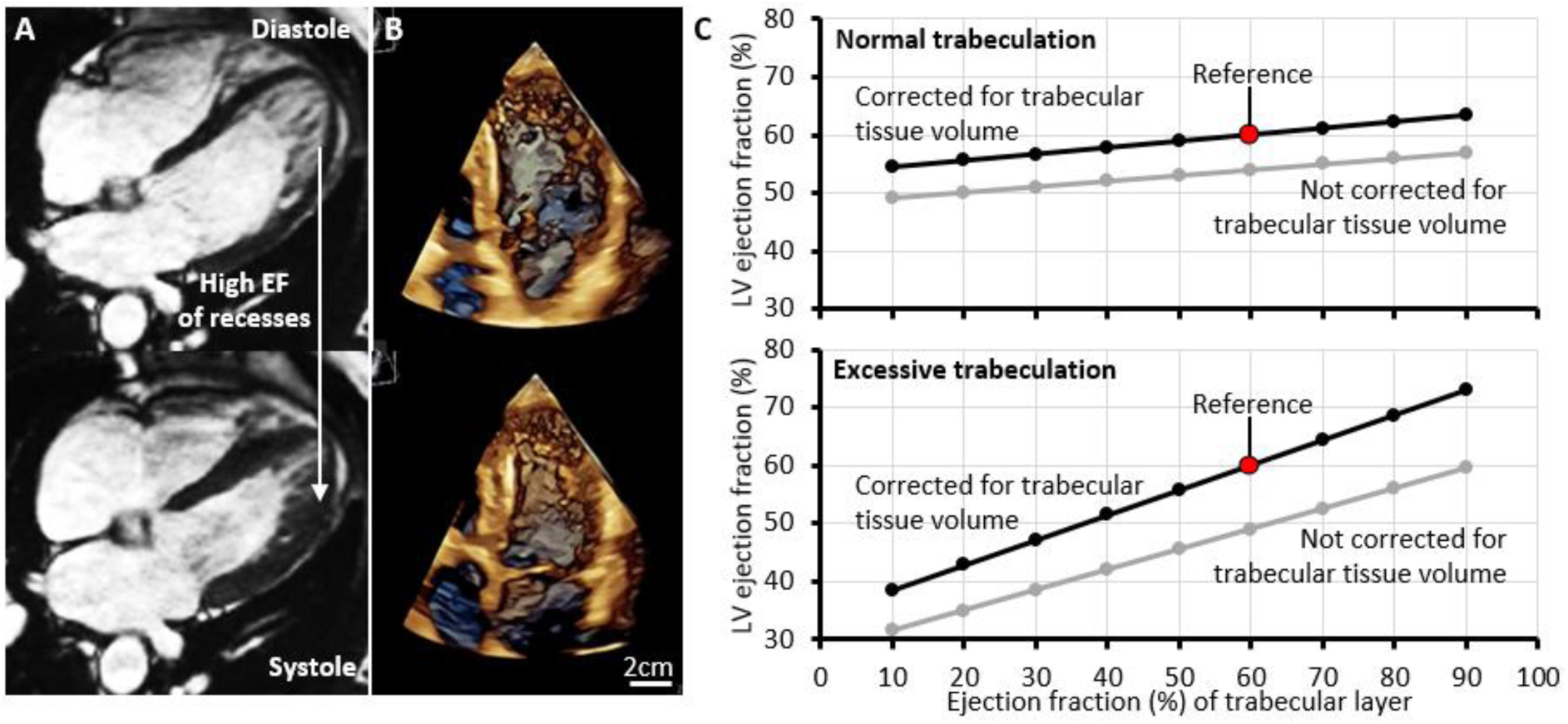
Impact of trabecular layer ejection fractions on total ejection fraction. **A**. CMR of a case of excessive trabeculation with a high ejection fraction of the trabecular layer. **B**. Same case as in A, visualized with 3D rendering of echocardiography, showing substantial compression of the intertrabecular recesses. **C**. The greater the ejection fraction of the trabecular layer, the greater the ejection of the total ventricular cavity (while the central cavity ejection fraction is kept at 60). Although this effect is subtle in a setting of normal trabeculation, the effect can be substantial in a setting of excessive trabeculation.

**Supplementary Figure S5.**
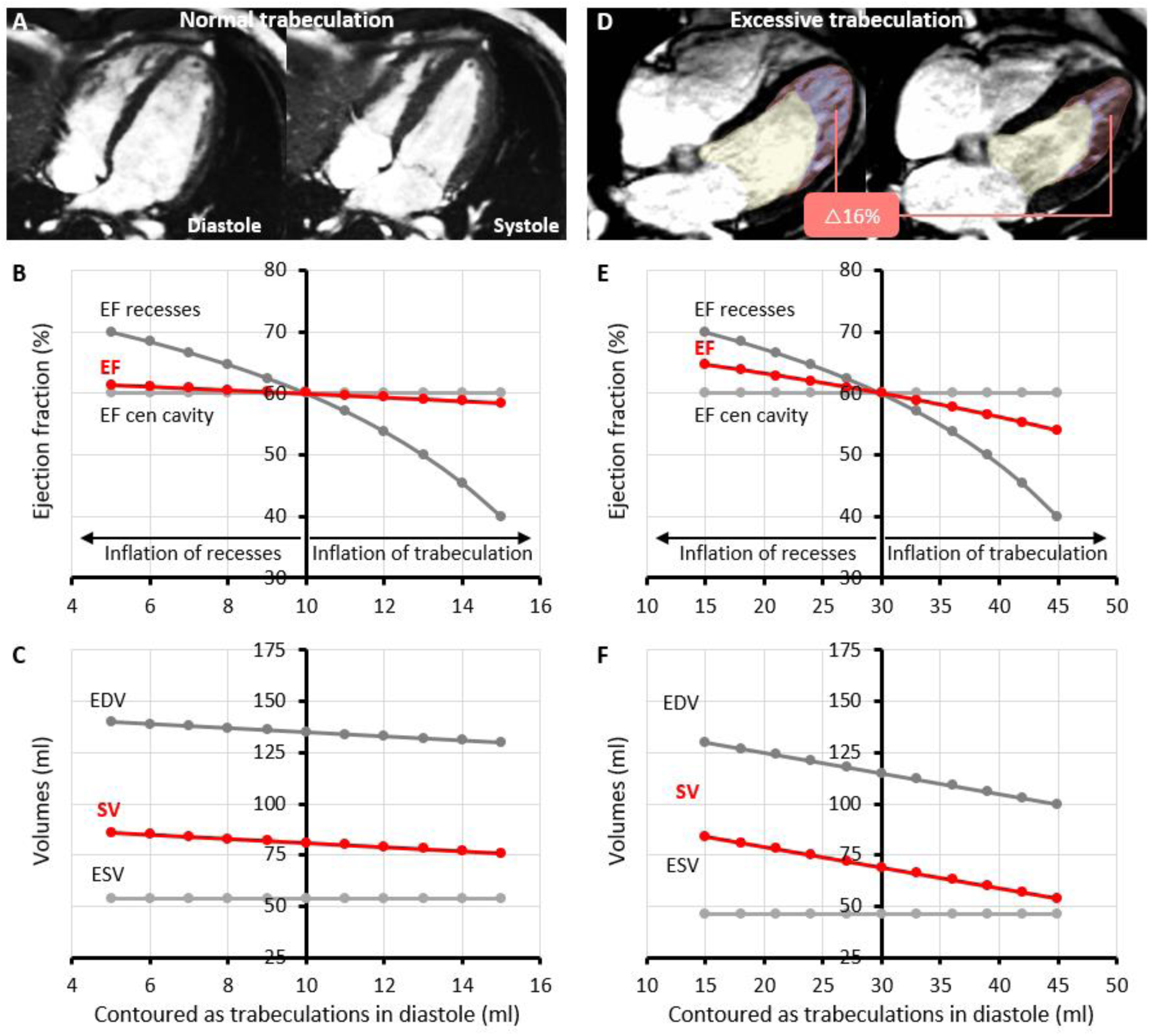
Theoretical calculations on the effect of discrepancies in trabecular myocardium measurements between diastole and systole. **A**-**C**. In a setting of normal trabeculation, in systole the trabeculations may be measured to comprise 10ml. Given this proportionally small volume, even substantially different labelling of the trabeculations in diastole, does not affect much the measured EF. **D**-**F**. In a setting of excessive trabeculation, in systole the trabeculations may be measured to comprise 30ml. Substantially different labelling of the trabeculations in diastole affects EDV, SV, and EF. EDV, end diastolic volume; ESV, end systolic volume; SV stroke volume; EF, ejection fraction.

## References

1. Dewey M, Siebes M, Kachelrieß M, Kofoed KF, Maurovich-Horvat P, Nikolaou K et al. Clinical quantitative cardiac imaging for the assessment of myocardial ischaemia. Nat Rev Cardiol 2020;17:427–450.

2. de la Chica JA, Gómez-Talavera S, García-Ruiz JM, García-Lunar I, Oliva B, Fernández-Alvira JM et al. Association Between Left Ventricular Noncompaction and Vigorous Physical Activity. J Am Coll Cardiol 2020;76:1723–1733.

3. Sigvardsen PE, Fuchs A, Kühl JT, Afzal S, Køber L, Nordestgaard BG et al. Left ventricular trabeculation and major adverse cardiovascular events: The Copenhagen General Population Study. Eur Heart J Cardiovasc Imaging 2021;22:67–74.

4. Woodbridge SP, Aung N, Paiva JM, Sanghvi MM, Zemrak F, Fung K et al. Physical activity and left ventricular trabeculation in the UK Biobank community-based cohort study. Heart 2019;105:990–998.

5. Polacin M, Károlyi M, Wilzeck V, Eberhard M, Gotschy A, Alkadhi H, Kozerke S, Manka R. Three-dimensional Whole-Heart Cardiac MRI Sequence for Measuring Trabeculation in Left Ventricular Noncompaction. Radiol Cardiothorac Imaging 2022;4:e220109.

6. Riekerk HCE, Coolen BF, Strijkers GJ, van der Wal AC, Petersen SE, Sheppard MN et al. Higher spatial resolution improves the interpretation of the extent of ventricular trabeculation. J Anat 2022;240:357–375.

7. Lang RM, Badano LP, Victor MA, Afilalo J, Armstrong A, Ernande L et al. Recommendations for cardiac chamber quantification by echocardiography in adults: An update from the American Society of Echocardiography and the European Association of Cardiovascular Imaging. J Am Soc Echocardiogr 2015;28:1–39.e14.

8. Davies RH, Augusto JB, Bhuva A, Xue H, Treibel TA, Ye Y et al. Precision measurement of cardiac structure and function in cardiovascular magnetic resonance using machine learning. J Cardiovasc Magn Reson 2022;24:16.

9. Choi Y, Kim SM, Lee S-C, Chang S-A, Jang SY, Choe YH. Quantification of left ventricular trabeculae using cardiovascular magnetic resonance for the diagnosis of left ventricular non-compaction: evaluation of trabecular volume and refined semi-quantitative criteria. J Cardiovasc Magn Reson 2016;18:24.

10. Dreisbach JG, Mathur S, Houbois CP, Oechslin E, Ross H, Hanneman K et al. Cardiovascular magnetic resonance based diagnosis of left ventricular non-compaction cardiomyopathy: impact of cine bSSFP strain analysis. J Cardiovasc Magn Reson 2020;22:9.

11. Jacquier A, Thuny F, Jop B, Giorgi R, Cohen F, Gaubert JY et al. Measurement of trabeculated left ventricular mass using cardiac magnetic resonance imaging in the diagnosis of left ventricular non-compaction. Eur Heart J 2010;31:1098–1104.

12. Stacey RB, Andersen MM, St. Clair M, Hundley WG, Thohan V. Comparison of Systolic and Diastolic Criteria for Isolated LV Noncompaction in CMR. JACC Cardiovasc Imaging 2013;6:931– 940.

13. Grothoff M, Pachowsky M, Hoffmann J, Posch M, Klaassen S, Lehmkuhl L et al. Value of cardiovascular MR in diagnosing left ventricular non-compaction cardiomyopathy and in discriminating between other cardiomyopathies. Eur Radiol 2012;22:2699–2709.

14. Jaspers K, Freling HG, Van Wijk K, Romijn EI, Greuter MJW, Willems TP. Improving the reproducibility of MR-derived left ventricular volume and function measurements with a semi-automatic threshold-based segmentation algorithm. Int J Cardiovasc Imaging 2013;29:617– 623.

15. Luu JM, Gebhard C, Ramasundarahettige C, Desai D, Schulze K, Marcotte F et al. Normal sex and age-specific parameters in a multi-ethnic population: a cardiovascular magnetic resonance study of the Canadian Alliance for Healthy Hearts and Minds cohort. J Cardiovasc Magn. Reson 2022;24:2.

16. Positano V, Meloni A, Macaione F, Santarelli MF, Pistoia L, Barison A et al. Non-compact myocardium assessment by cardiac magnetic resonance: dependence on image analysis method. Int J Cardiovasc Imaging 2018;34:1227–1238.

17. Thut T, Valsangiacomo Büchel E, Geiger J, Kellenberger CJ, Rücker B, Burkhardt BEU. Signal Thresholding Segmentation of Ventricular Volumes in Young Patients with Various Diseases— Can We Trust the Numbers? Diagnostics. 2023;13:180.

18. Petersen SE, Jensen B, Aung N, Friedrich MG, McMahon CJ, Mohiddin SA et al. Excessive Trabeculation of the Left Ventricle: JACC: Cardiovascular Imaging Expert Panel Paper. JACC Cardiovasc Imaging 2023;16:408–425.

19. Riffel JH, Andre F, Maertens M, Rost F, Keller MGP, Giusca S et al. Fast assessment of long axis strain with standard cardiovascular magnetic resonance: A validation study of a novel parameter with reference values. J Cardiovasc Magn Reson 2015;17:1–9.

20. Barskiy V. Arrhythmogenic right ventricular cardiomyopathy. Case study, Radiopaedia.org. 10.53347/rID-69431. Accessed 18 Feb 2023.

21. Barskiy V. Non-compaction of the left ventricle. Case study, Radiopaedia.org. 10.53347/rID-69436. Accessed 18 Feb 2023.

22. Kaplan-List K. Secundum atrial septal defect. Case study, Radiopaedia.org. 10.53347/rID-42823. Accessed 18 Feb 2023.

23. Keshavamurthy J. Incidental infundibular pulmonary stenosis and left sided superior vena cava. Case study, Radiopaedia.org. 10.53347/rID-44210. Accessed 18 Feb 2023.

24. Luijkx T. Left ventricular non-compaction. Case study, Radiopaedia.org. 10.53347/rID-39933. Accessed 18 Feb 2023.

25. Luijkx T. Physiological cardiac adaptation to exercise. Case study, Radiopaedia.org. 10.53347/rID-39934. Accessed 18 Feb 2023.

26. O’Rourke R. Endocardial fibroelastosis. Case study, Radiopaedia.org. 10.53347/rID-86346. Accessed 18 Feb 2023.

27. Sheehy N. Apical hypertrophic cardiomyopathy. Case study, Radiopaedia.org. 10.53347/rID-79346. Accessed 18 Feb 2023.

28. Tigges S. Left ventricular non-compaction. Case study, Radiopaedia.org. 10.53347/rID-158103. Accessed 18 Feb 2023.

29. Yarmola I. Dilated cardiomyopathy with non-compaction of the left ventricle. Case study, Radiopaedia.org. 10.53347/rID-69559. Accessed 18 Feb 2023.

30. Yousef ZR, Foley PWX, Khadjooi K, Chalil S, Sandman H, Mohammed NUH et al. Left ventricular non-compaction: Clinical features and cardiovascular magnetic resonance imaging. BMC Cardiovasc Disord 2009;9:37.

31. Martins E, Pinho T, Carpenter S, Leite S, Garcia R, Madureira A et al. Histopathological evidence of Fabry disease in a female patient with left ventricular noncompaction. Rev Port Cardiol 2014;33:565.e1–565.e6.

32. Chan VS, Chan CW, Cheung SC. Cardiac magnetic resonance imaging in the diagnosis of biventricular non-compaction in a young but failing heart. Hong Kong Med J 2019;25:**330**.e1–330.e2.

33. Paluszkiewicz J, Milting H, Kałużna-Oleksy M, Pyda M, Janus M, Körperich H et al. Left Ventricular Non-Compaction Cardiomyopathy-Still More Questions than Answers. J Clin Med 2022;11:4135.

34. Takamatsu M, Kamohara K, Sato M, Koga Y. Effect of Noncompacted Myocardial Resection on Isolated Left Ventricular Noncompaction. Ann Thorac Surg 2020;110:e387–e389.

35. Faber JW, Wüst RCI, Dierx I, Hummelink JA, Kuster DWD, Nollet E et al. Equal force generation potential of trabecular and compact wall ventricular cardiomyocytes. iScience 2022;25:105393.

36. Litviňuková M, Talavera-López C, Maatz H, Reichart D, Worth CL, Lindberg EL et al. Cells of the adult human heart. Nature 2020;588:466–472.

37. McDonagh TA, Metra M, Adamo M, Gardner RS, Baumbach A, Böhm M et al. 2021 ESC Guidelines for the diagnosis and treatment of acute and chronic heart failure. Eur Heart J 2021;**42**:3599–3726.

38. Zemrak F, Ahlman MA, Captur G, Mohiddin SA, Kawel-Boehm N, Prince MR et al. The Relationship of Left Ventricular Trabeculation to Ventricular Function and Structure Over a 9.5- Year Follow-Up. J Am Coll Cardiol 2014;64:1971–1980.

39. Williams CJA, Greunz EM, Ringgaard S, Hansen K, Bertelsen MF, Wang T. Magnetic Resonance Imaging (MRI) reveals high cardiac ejection fractions in red-footed tortoises (Chelonoidis carbonarius). J Exp Biol 2019;222:18

